# Lipidome- and genome-wide study to understand sex differences in circulatory lipids

**DOI:** 10.1101/2022.05.30.22275704

**Authors:** Rubina Tabassum, Sanni Ruotsalainen, Linda Ottensmann, Mathias J. Gerl, Christian Klose, Taru Tukiainen, Matti Pirinen, Kai Simons, Elisabeth Widén, Samuli Ripatti

## Abstract

Despite well-recognized difference in the atherosclerotic cardiovascular disease (ASCVD) risk between men and women, sex differences in risk factors and sex specific mechanisms in the pathophysiology of ASCVD remain poorly understood. Lipid metabolism plays a central role in the development of ASCVD. Understanding sex differences in lipids and their genetic determinants could provide mechanistic insights into sex differences in ASCVD and aid in precise risk assessment. Thus, we examined sex differences in plasma levels of 179 lipid species from 7,266 participants and performed sex-stratified genome-wide association studies (GWAS) to evaluate contribution of genetic factors in sex differences. We sought for replication using independent data from 2,045 participants. Significant sex differences in levels of 141 lipid species were observed (P<7.0×10^−4^). Interestingly, 121 lipid species showed significant age-sex interactions with opposite age-related changes in 39 lipid species. In general, most of the cholesteryl esters, ceramides, lysophospholipids and glycerides were higher in 45-50-year-old men compared with women of same age, but the sex-differences narrowed down or reversed with age. We did not observe any major differences in genetic effect in the sex stratified GWAS which suggests that common genetic variants do not have a major role in sex differences in lipidome. In conclusion, our study provides a comprehensive view of sex differences in circulatory lipids pointing to potential sex differences in lipid metabolism, highlighting need for sex- and age-specific prevention strategies.

## Introduction

Although atherosclerotic cardiovascular disease (ASCVD) is the leading cause of death both among men and women worldwide [1], there are substantial sex differences in the prevalence and burden of its manifestations [2, 3]. Despite many efforts, ASCVD remains understudied, under-recognized, underdiagnosed, and undertreated in women [4]. Limited understanding of sex differences in etiology and clinical presentations of ASCVD often leads to misdiagnosis in women resulting in higher disease burden and mortality among women compared with men [4]. This emphasizes an urgent need for unravelling underlying biological mechanisms that contribute to sex differences in ASCVD pathophysiology to develop sex specific strategies for early detection and prevention.

Plasma lipids are well-established heritable risk factors for ASCVD [5] and are routinely monitored to assess its clinical risk [6, 7]. Sex differences in plasma levels of total cholesterol, triglycerides, high-density lipoprotein cholesterol (HDL-C) and low-density lipoprotein cholesterol (LDL-C) (referred henceforth as traditional lipids) have been recognized [8-9]. However, understanding of sex differences in detailed lipidomes is limited and less explored [10-12]. Several studies have demonstrated potential of lipidomics that allow simultaneous measurements of hundreds of lipid species or subspecies in understanding ASCVD and risk prediction beyond traditional lipids [13-17]. Integration of lipidomics with genomics has also provided new insights into genetic regulation of lipid metabolism and ASCVD pathophysiology [18-23]. Strong influence of genetic variants such as *FADS1-2-3, LDLR, PCSK9* in maintenance of lipid homeostasis calls for a comprehensive evaluation of potential role of genetic mechanisms in sex differences in lipid metabolism. Thus, stronger consideration should be given to understand sex differences in lipidome and their genetic determinants.

In this study, we examined sex differences in human plasma lipidome in a large dataset comprising of 179 lipid species measured by shotgun lipidomics from 7,266 participants from the GeneRISK cohort. We further performed sex-stratified genome-wide association analysis for all the lipid species to evaluate contribution of genetic factors in sex differences in lipidomes. We sought for replication of the findings in an independent dataset comprising of 169 lipid species from 2,045 participants measured by the same lipidomics platform. Our results show that lipidome exhibit significant age-dependent differences between men and women pointing to potential sex differences in lipid metabolism and highlights the need for sex- and age-stratified analyses in lipidome studies. Moreover, sex-stratified genome-wide association analyses did not suggest a major role of common genetic variants in sex differences in lipidome. Overall, our study emphasizes the need for sex-specific prevention and management of ASCVD risk.

## Results

After quality control, lipidomics data in GeneRISK cohort comprised of 179 lipid species from 13 lipid classes from 7,266 individuals including 2,624 men and 4,642 women. The basic clinical characteristics of the study population are provided in S1 Table. The details of the lipid species included in the study and their mean plasma levels are provided in (S1 Fig and S2 Table). Men and women participants had similar age distribution ranging from 45-66 years, with mean age of 55.9 (±5.9) and 55.7 (±5.7) years, respectively. Consistent with the known sex differences in ASCVD risk, women participants had relatively lower estimated 10-year ASCVD risk compared with men (Fig 1A). Men had significantly higher levels HDL-C, LDL-C, TG, ApoA1 and ApoB compared with that of women (S3 Table). However, levels of TC, LDL-C and TGs showed age-dependent sex differences (age-sex interaction term P < 7.0×10^−4^), resulting in either reversed or abrogated differences in older age-groups (Fig 1B). Similar age-dependent sex differences in traditional lipids were observed in the UK Biobank dataset (S2 Fig and S3 Table).

**Fig 1:**
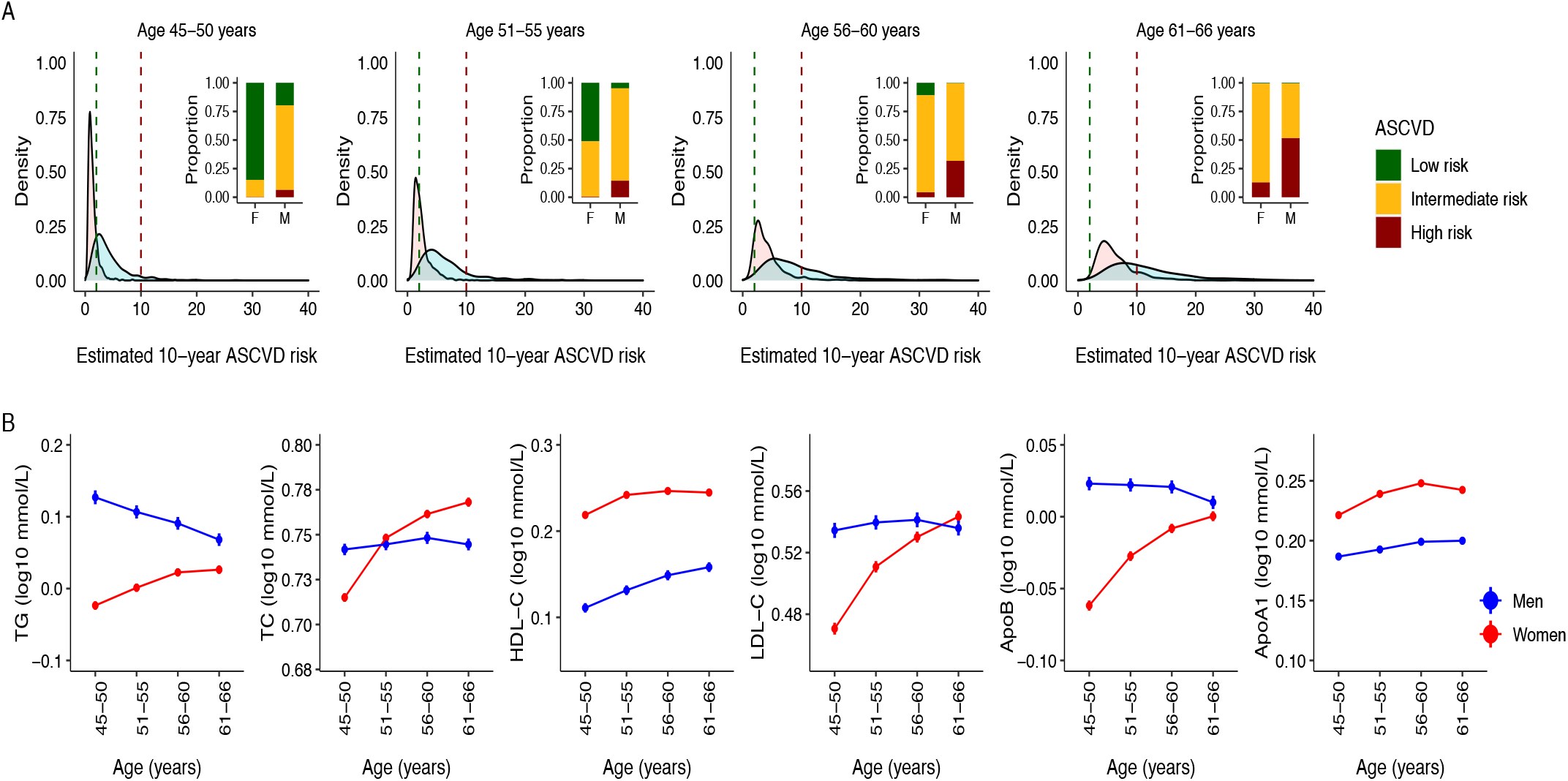
Sex differences in ASCVD risk and traditional plasma lipids. (A) Distributions of 10-year ASCVD risk estimated based on traditional risk factors in men (blue) and women (pink) at 5-year interval are shown in the density plots. The vertical green and red lines on density plots mark the low (< 2%) and high (> 10%) 10-year estimated risk for ASCVD, respectively. The bar plot inserts in each density plot show the proportion of men (M) and women (F) with low risk (< 2%), intermediate risk (2-10%) and high risk (> 10%) for ASCVD in the respective age-group. Individuals with preexisting medical conditions (N=795) were not included in data used for these plots. (B) Age-related trends for traditional lipids in the GeneRISK cohort. Mean levels after log10 transformation and standard errors of total cholesterol, triglycerides, HDL-C, LDL-C, Apolipoprotein B and Apolipoprotein A1 at 5-year interval are plotted for men (blue) and women (red). Individuals with lipid lowering medications were excluded before calculating the mean levels of lipids for this analysis.

### Age and sex interactions in lipid species levels

Of 179 lipid species, significant differences in plasma levels of 141 lipid species between men and women were observed (Fig 2A). However, as descriptive analysis of traditional lipids suggested significant effect of age and sex interaction in circulating lipid levels, we further evaluated for age-sex interactions in individual lipid species. Significant age-sex interactions for 121 lipid species (interaction term P < 7.0×10^−4^) were found in models including covariates and age-sex interaction term (S4 Table). A total of 116 out of these 121 lipid species also showed significant sex-heterogeneity in their effect sizes for association with age (P_het_ < 7.0×10^−4^) (Fig 2B, S5 Table). Effect of age-sex interactions on lipidome is also evident from the heatmaps depicting 1-year change (in standard deviation units adjusted for covariates) in plasma levels of each lipid species in men and women relative to 45–50-year-old men and women respectively (S3 Fig, S6 Table). Among men, age was associated with decrease in levels of 72 lipid species and increase in levels of 10 lipid species (S5 Table, S4 Fig). On the contrary, age was positively associated with 108 lipids and negatively associated only with 3 lipid species in women. Interestingly, of the 51 lipid species that were significantly associated with age both in men and women, 39 lipid species showed opposite age-related trends in men and women. Sensitivity analyses after excluding individuals with lipid-lowering medications provided similar results (S5 Table).

**Fig 2:**
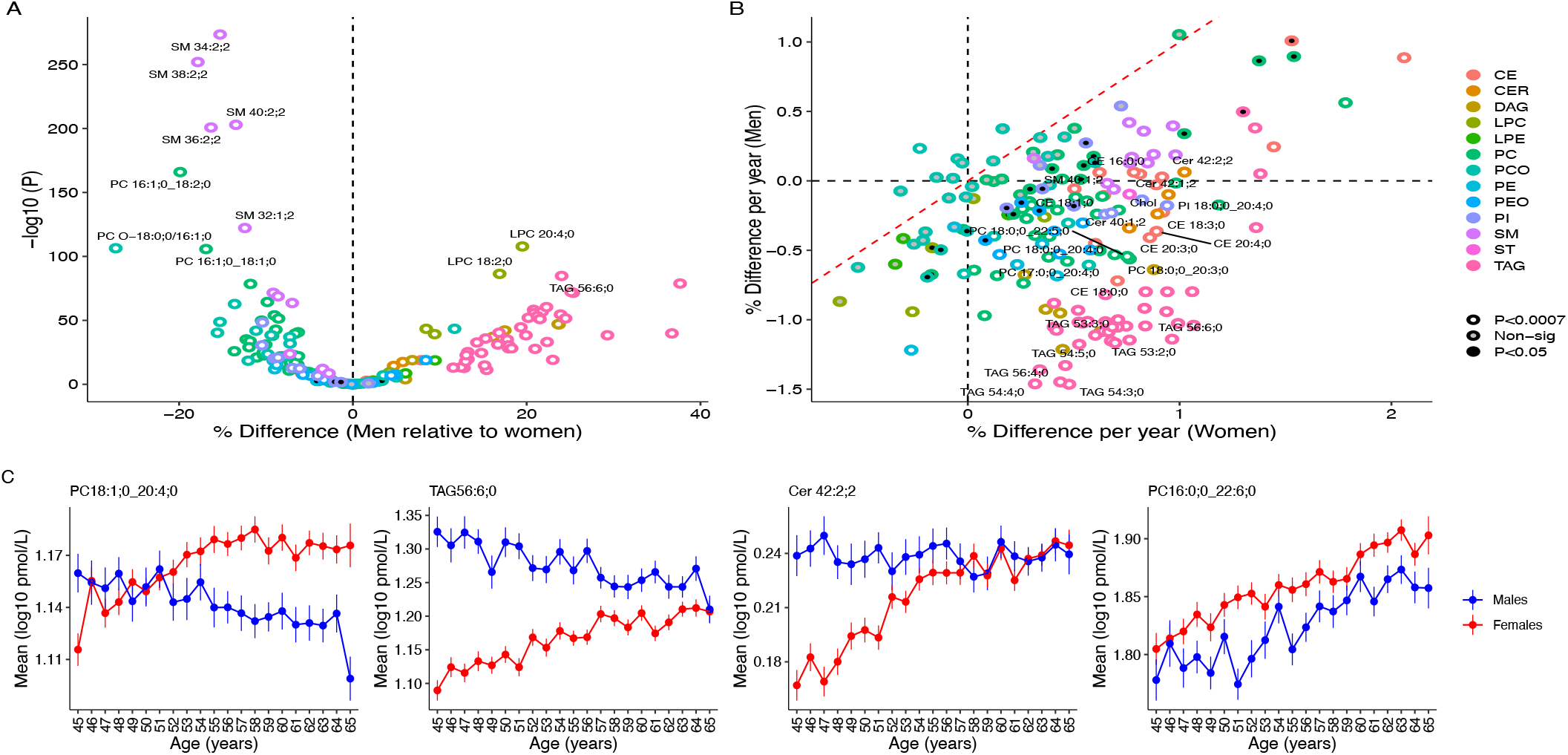
Age and sex interactions in the levels of circulatory lipids. (A) Association of 179 lipid species with sex. Scatter plot shows percent differences in lipid levels between men and women in full dataset on x-axis and the corresponding P values on y-axis. Positive difference means higher level in men and negative difference means higher level in women. (B) Heterogeneity in association of lipid species with age between men and women. Scatter plot shows percent difference per year in women on x-axis and percent difference per year in men on y-axis. Lipid species with significant heterogeneity (P_het_ < 7.0×10^−4^) are filled with white color and colored by lipid class. (C) Age-related trends for representative lipid species in men and women illustrating effect of age on sex differences. Mean plasma levels with standard error for 1-year interval are plotted for men (blue) and women (red). PC 18:1;0_20:4;0 - Opposite trends in men and women leading to increased sex differences with age; TAG 56:6;0 - Opposite trends in men and women for resulting in narrowing of sex differences; Cer 42:2;2 – Association with age only in women resulting in narrowing of sex difference; PC 16:0;0_22:6;0 - Similar age-related trends for in men and women with no effect heterogeneity.

Consistent age-sex interactions were also observed in replication analysis that included 169 lipid species that matched with the GeneRISK lipidomics dataset (S7 Table). From 121 lipid species with significant age-sex interactions in the GeneRISK cohort, 113 lipid species were available in replication cohort, of which 88 lipid species had significant age-sex interactions (P < 0.05) (S4 Table). Moreover, of the 82 and 111 lipid species that were associated with age in men and women respectively in the GeneRISK, 67 and 52 lipid species respectively (out of 79 and 104 respectively available in replication cohort) were validated in replication cohort at (P < 0.05) (S8 Table, S5 Fig). Overall, most of the age-associated lipid species in the GeneRISK had similar effect sizes in the replication cohort (r^2^ in men = 0.84; r^2^ in women = 0.63) (S6 Fig).

### Age-dependent sex differences in lipid species levels

As we observed significant age-sex interactions in the lipidome, we present sex differences in lipid species at 5-year intervals (S9 Table). While levels of 81 lipid species including triacylglycerides (TAGs), diacylglycerides (DAGs), cholesteryl esters (CEs), ceramides (Cers), lysophospholipids (LPCs) were higher among men of 45-50 years of age, decline in their levels with age resulted in significantly lower levels of most lipid species (N=85) in 61–66-year-old men compared with the women of same age-range. Effect of distinct age-related changes on sex differences in lipid levels are depicted for representative lipid species in Fig 2C and for all lipids in S7 Fig. Sex differences in TAGs and DAGs narrowed down with age, and finally resulted in similar TAGs and DAGs profiles in men and women in older age (Fig 3). On the other hand, sex differences in sphingomyelins (SMs) continued to increase with age; while sex differences in most of the phospholipids and CEs either abrogated or reversed in direction with age, resulting in lower phospholipids and CEs profiles in older men compared with women of same age-group. Altogether, from the 109 lipid species that had significant sex differences in 45–50-year-old participants, 55 lipid species including TAGs, DAGs, and phospholipids with polyunsaturated fatty acids (PUFAs) did not remain significant in older participants while 11 lipid species reversed the gaps. Analysis in the replication cohort provided similar trends in age-dependent sex differences in most of the lipid species (S10 Table and S8 Fig). Strong correlation between effect sizes for association with sex in the GeneRISK and replication cohort at different age-groups was found (r^2^_45-50y_ = 0.94, r^2^_51-55y_ = 0.85, r^2^_56-60y_ = 0.85, r^2^_61-66y_ = 0.75) (S9 Fig).

**Fig 3:**
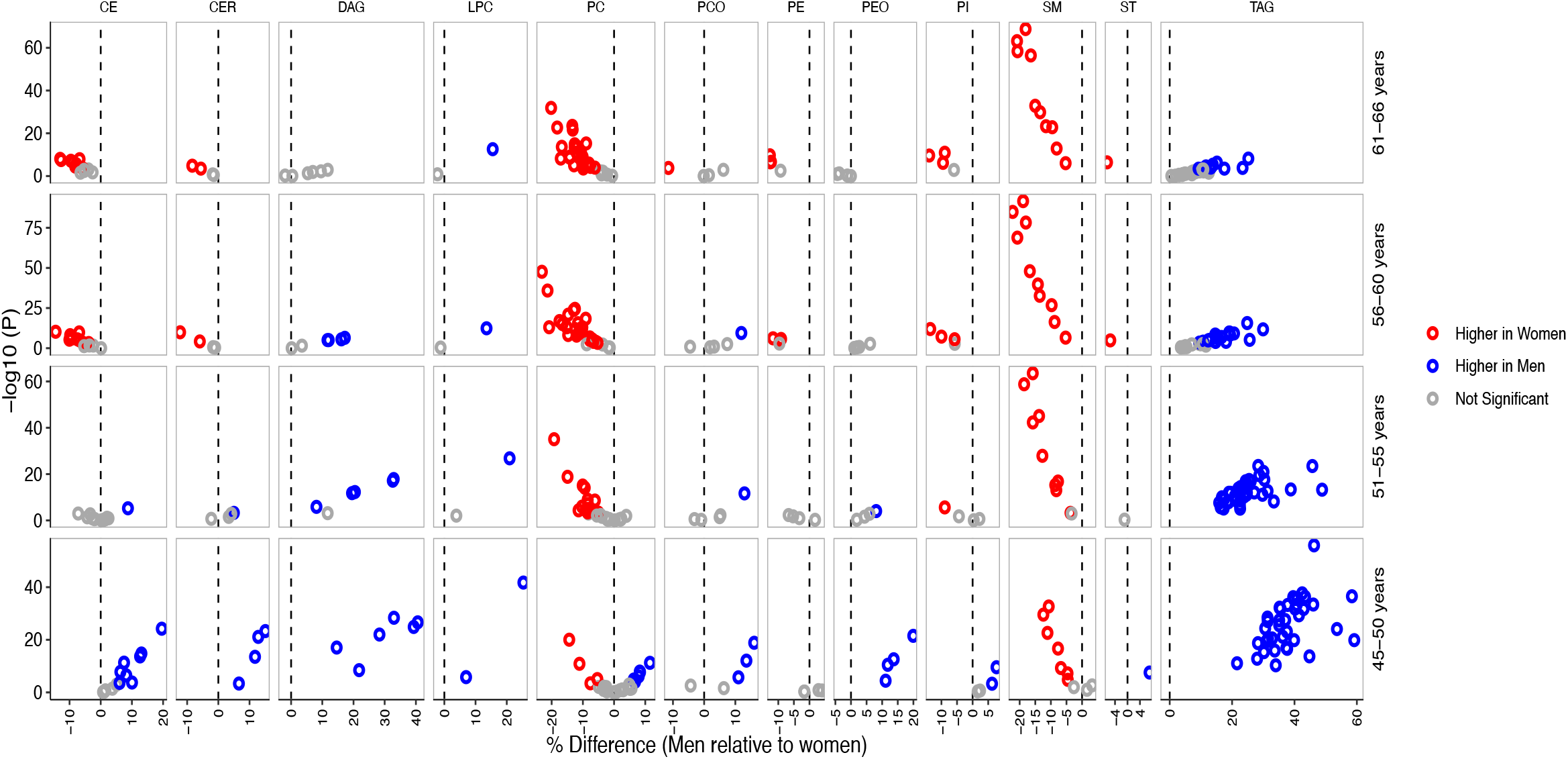
Age-dependent sex differences in plasma lipidome. Association of lipid species with sex in different age groups are shown with lipid species being grouped by lipid classes. The x-axes show percent differences in men relative to women and the corresponding P values on y axes. Positive difference represents higher lipid level in men (blue color) and negative difference represents higher lipid level in women (red color) after multiple testing correction (P < 7.0×10^−4^). Only the lipid species with significant age and sex interactions are plotted here for clarity.

As menopause has previously been suggested to have effect on lipid levels, we performed exploratory analyses to investigate effect of menopause status on the observed age-dependent sex differences in lipidome. Only two lipid species Cer 42:2;2 and SM 38:2;2 showed significant association with menopause status (P < 7.0×10^−4^) (S10 Fig, S11 Table). No significant interaction between age and menopause was found for any of the lipid species (S12 Table). Moreover, comparison of differences in lipid levels between men compared with premenopausal women and men compared with postmenopausal women revealed effect heterogeneity only in 17 lipid species (P_het_ < 7.0×10^−4^) (S13 Table, S10 Fig). Overall, our results do not support a major effect of menopause on the plasma levels of lipid species analyzed here and their sex differences.

### Effect of genetic factors on sex differences in lipidome

To evaluate if the observed sex differences in lipidome are due to differential effect of genetic variants in men and women, we conducted two types of genome-wide searches: (a) genome-wide heterogeneity scan and (b) sex-specific association scans. In the genome-wide heterogeneity scan, variants with P_het_ < 7.0×10^−10^ (5.0×10^−8^/70 PCs explaining >90% variance in lipidomes) were defined as sex-dimorphic variants. Based on this criteria, sex-heterogeneity was found at rs7760319 in the intron of *SASH1* for PC 17:0;0_20:4;0 (P_het_ = 1.9×10^−10^) (Fig 4A), that was nominally associated with the lipid with opposite effects in men (ß_men_ = - 0.13±0.03, P_men_= 3.7×10^−6^) and women (ß_women_ = 0.10±0.02, P_women_ = 1.2×10^−5^) in the sex-stratified GWAS. However, the signal was not validated in the replication cohort (ß_men_ = - 0.015±0.047, P_men_ = 0.75; ß_women_ = 0.01±0.045, P_women_ = 0.84). List of all the associations with P_het_ < 5.0×10^−8^ is provided in S14 Table.

**Fig 4:**
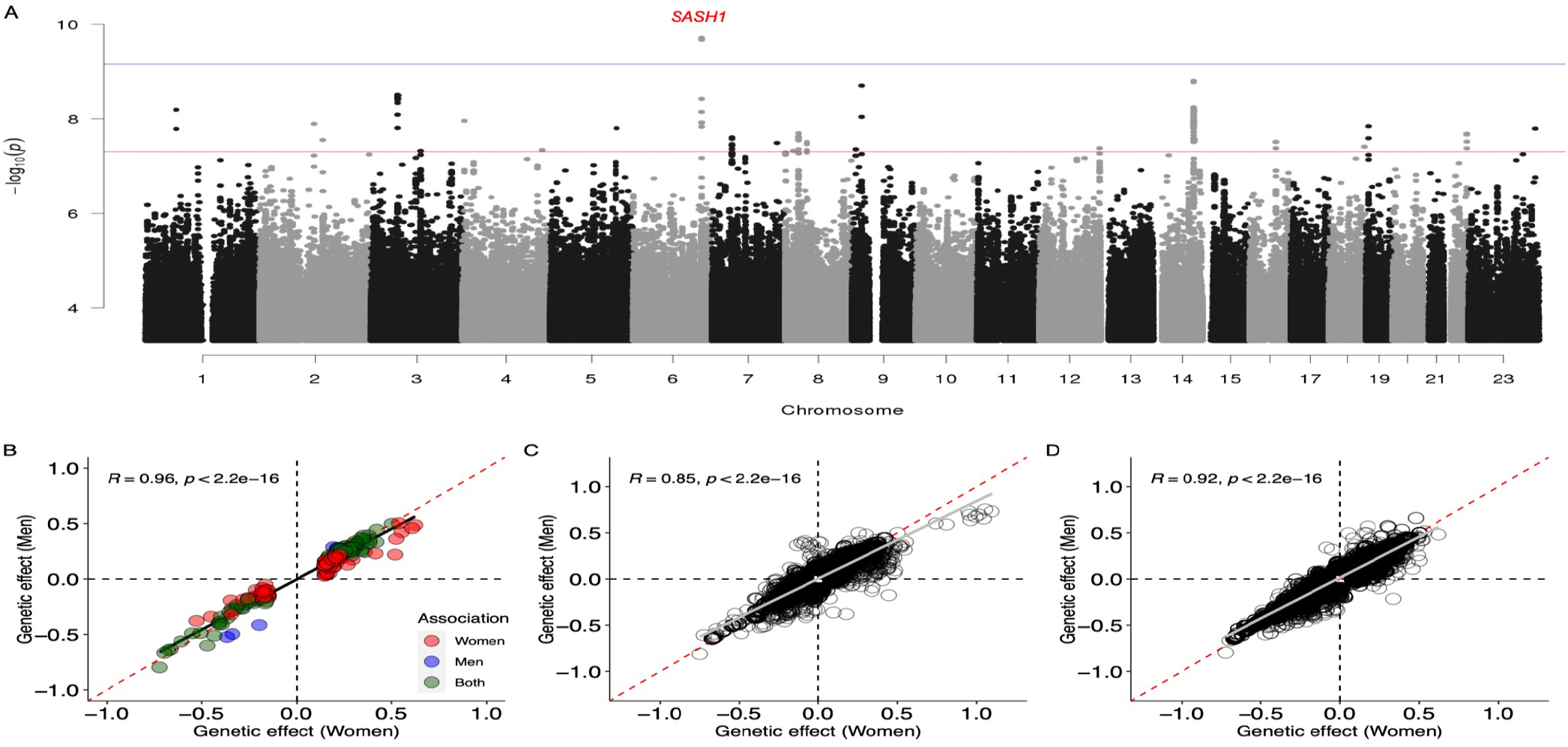
Effect of genetic heterogeneity on sex differences in lipidome. (A) Genome-wide heterogeneity in genetic effect on lipidome. Manhattan plot shows -log10 P_het_ on y-axis for all the lipid species and genetic variants on x-axis arranged by chromosome position on different chromosomes. Red and blue horizontal lines represent P value threshold for genome-wide significance and after multiple testing correction. (B) Heterogeneity in effect of variants identified in sex-stratified GWAS. Scatter plot shows comparison of beta estimates for 193 unique locus-lipid associations with P < 7.0×10^−10^ identified in sex-stratified GWAS. Blue and red colored points represent associations reaching threshold for multiple testing correction (P < 7.0×10^−10^) in GWASs for men and women respectively, while green colored points represent associations reaching threshold for multiple testing correction in GWASs for both men and women. (C) Heterogeneity in effect of variants identified in previous GWAS for traditional lipids. (D) Heterogeneity in effect of variants identified in previous GWAS for lipidome. The scatter plots in (C) and (D) shows genetic effect of known lipid loci in men (y-axis) and women (x-axis) on lipidome. Associations with P < 7.0×10^−4^ are plotted.

In the sex-specific association scans, variants with significant associations (P < 7.0×10^−10^) either in men or in women in sex-stratified GWAS were selected and tested for heterogeneity in their effects between men and women. In male-specific GWAS, 5,828 variant-lipid pair associations were found, while female-specific GWAS identified 13,392 variant-lipid pair associations (S15 and S16 Tables). In total, 193 unique locus-lipid associations involving 26 independent genomic loci and 127 lipid species either in men or in women were identified and tested for differences in their effect estimates between men and women. Using P_het_ < 2.5×10^−4^ (0.05/193) as the threshold, none of the tested variants showed significant heterogeneity (S17 Table), rather the effect sizes for these associations were quite similar and highly correlated in men and women (Fig 4B). We further tested for heterogeneity in effects of the genetic variants identified in previous GWASs for traditional lipids and lipidome (S18 and S19 Tables), which also suggested that effect estimates of the known lipid variants are similar in men and women as shown in Fig 4C and 4D.

We further explored for the heterogeneity in the genomic regions with genes encoding enzymes with desaturase and elongase activities - *FADS1-2-3, SCD1, ELOVL5* and *ELOVL6* for association with lipid indices representing efficiency of respective enzymes. The lipid indices used in the analysis and their calculations are provided in S20 Table. It is to be noted that such indices and ratios have been applied previously to free fatty acids but their validity for complex lipids are not known. Though no strong evidence for heterogeneity at these regions was found, nominal heterogeneity at *FADS1-2-3* and *SCD1* regions were observed and are listed in S21 Table. Noteworthy, at *FADS1-2-3* region, a non-synonymous variant rs35723406 in *TKFC* gene showed effect heterogeneity between men and women for Δ6 desaturase in TAG (D6D in TAG) (P_het_ = 6.2×10^−6^) (S11 Fig), which might be of interest for further investigation.

## Discussion

This study presents a comprehensive characterization of sex differences in circulatory lipids using high coverage lipidome profiles and effect of age and genetic factors on differences in lipidome between men and women. The results show significant sex differences in the lipidome; and demonstrate that age has distinct effect on lipidomes in men and women. In general, most of the CEs, Cers, LPCs, TAGs and DAGs were higher in men at 45-50 years of age, but these sex differences tend to narrow down or reverse with age. Our results did not provide support for major roles of menopause and common genetic factors in sex differences in plasma lipidome.

Age-specific sex differences in circulatory lipids could have important implications in risk assessment and precision medicine for cardiometabolic diseases. Sphingolipids including Cers and SMs have emerged as promising new diagnostic or prognostic markers for CVD with potential clinical utility. CERT score based on Cer(d18:1/16:0), Cer(d18:1/18:0) and Cer(d18:1/24:1) and their ratios with Cer(d18:1/24:0) is shown to predict cardiovascular death in patients with stable CAD and acute coronary syndromes beyond LDL-C [24]. Another risk score termed as sphingolipid-inclusive CAD (SIC) risk score that includes SMs in addition to Cers has been proposed as a strong predictor of CVD [15]. In this study, we found that Cer(d18:1/24:0) (measured as Cer 42:1;2) and Cer(d18:1/24:1) (measured as Cer 42:2;2) are significantly higher in 45–50-year-old men compared with women but their plasma levels become similar in men and women in older age-groups (>55 years) due to increase in their levels with age among women. A previous study also reported higher levels of ceramides in women aged >45 years compared with men [25]. However, we did not find significant increase in ceramide levels in men with age as reported earlier [10, 25]. Similarly, SMs levels were higher among women compared with men and their levels increased with age in women, as also reported previously [10, 26]. These observations of increased levels of Cers and SMs in older women are consistent with the known increase in CVD risk with age in women. Our results also point that sex differences in sphingolipids need to be considered in Cers and SMs based prediction scores.

In addition to sphingolipids, many lipid species including PCs and TAGs have been identified as risk factors for CVD. It has been consistently demonstrated that phospholipids with saturated and monounsaturated fatty acyl-chains are positively associated with CVD risk, while polyunsaturated phospholipids are inversely associated with CVD risk [17]. On similar lines, we found that while most of PCs increase with age in women, among men, mainly PCs with PUFAs change with age and most of the PCs with saturated and monounsaturated FA were not associated with age. Similarly, TAGs have opposite age-related trends in men and women, with significantly higher levels of most of TAGs in men until 60 years of age. All these observations further emphasize the importance of considering sex differences in lipids in ASCVD risk assessment and sex-specific prevention strategies.

Our findings also suggest potential sex differences in phospholipids and glycerolipids metabolism. Opposite age-related trends for TAGs in men and women point to the sex-specific regulation of triglyceride metabolism with age. Our study also suggests sex-dimorphism in PUFA metabolic pathway. We found that CEs and PCs with C20:3 and C20:4 fatty acids have opposite age-related trends in men and women (increase in women and decrease in men), but those with C20:5 and C22:6 had similar age-related trend i.e., they increase with age in both sexes. Our results also suggest that age and sex do not have substantial effect on lipid species with less unsaturated fatty acids and may have stronger regulation by other endogenous factors. These findings call for further studies to understand the underlying mechanisms.

Menopause has been reported to contribute to the increase in CVD risk with age in women and is accompanied by unfavorable changes in CVD risk factors including lipid profiles [27-29]. However, the impact of menopause and depleted endogenous estrogen levels distinct from that of advancing age remains controversial [30, 31]. An alternate hypothesis has emerged that increased premenopausal cardiovascular risk promotes early menopause has been proposed and supported by a few studies [32-34]. This implies that the direction of causality in the relationship between menopause and CVD is unclear. There have been inconsistencies in the association of lipids with menopause status also. Higher levels of TC, LDL-C, TG and ApoB and subfraction of lipoproteins and their lipid contents have been associated with menopause [35]. Beyene et al. also reported higher levels of PCs, PIs, SMs, PEs, Cers and TAGs in postmenopausal women [10]. Contrary to these observations, a recent study does not support a major role of menopause in lipid level changes [36]. Our study also did not find substantial effect of menopause status on sex differences in lipidome, although the reduced sample size in the menopause related analyses limit statistical power. Moreover, as age and menopause status are highly collinear variables, it is difficult to tease out the independent effect of age and menopause on outcome variables. Further investigation in this regard is important to understand the role of menopause in modulating lipid levels.

Contribution of genetic factors in endogenous regulation of lipid metabolism is well-recognized. Despite the known and expected differences in the lipid levels, men and women are typically analyzed together using sex as a covariate to account for potential sex differences in the lipid levels. Sex-combined analyses are under-powered to detect significant associations if the effects are in opposite directions. Though sex-heterogeneity in effect of a few lipid loci such as *LPL, APOE*, and *KLF* on traditional lipids has been reported [37, 38], there has been no effort to systematically evaluate differential effects of genetic variants on plasma lipidome in men and women. To our knowledge, our study presents the first comprehensive investigation of sex-dimorphism in genetic influences on plasma lipidome. Our observation that genetic factors have no major impact on sex differences in plasma levels of lipid species is in accordance with the findings from metabolomics-based study [11, 39, 40]. In a sex-specific GWAS study by Mittelstrass et al. that included many lipid species from PCs, LPCs, and SMs, no genome-wide significant differences in beta-estimates for SNPs for any of the lipid species was found [40]. Although, we acknowledge the challenge of limited statistical power for sex-stratified analyses, our finding of distinct age-related changes in the levels of lipid for males and females underlines that these differential profiles need to be accounted for to detect sex-dimorphic genetic influence. Given the sample size of the present study, we did not have sufficient statistical power to perform age-stratified GWAS, but a recent study highlighted difference in the influence of polygenic risk score for coronary heart disease on apolipoprotein in different age tertiles emphasizing the importance of age-stratified analysis [41]. Our study is the first effort towards understanding genetic mechanisms in sex differences in lipidome and we believe that it would pave the way for further studies in the direction.

Though the study is based on a large population-based cohort with a broad lipidome coverage, it is not without limitations. This is a cross-sectional study and the findings of effect of age on lipidome need to be replicated in longitudinal studies with long follow-up durations. Our study included participants aged 45-66 years. A wider range of age would be needed to provide more insights to the changes in lipidome profile in early adulthood and to get better picture of effect of menopause on lipidome. As suggested in our study, demographic and other cohort characteristics could affect the lipidome profiles, it is not clear if findings of this study could be generalized to other populations. However, it is to be noted that most of our findings are consistent or in accordance with the current understanding in the field. Furthermore, lipidomic profiles were measured in whole plasma, which does not provide information at the level of individual lipoprotein subclasses and limits our ability to gain detailed mechanistic insights. Further advances in lipidomics platforms might help to capture more comprehensive and complete lipidomic profiles, including the position of fatty acyl-chains in the glycerol backbone of TAGs and glycerophospholipids and detection of sphingosine-1-P species and several other species, that would allow to overcome these limitations.

In conclusion, our study reports considerable insights into the sex differences and age-related trends in circulatory lipids at molecular lipid species level. We show that men and women have distinct age-related changes in lipidome profiles that result in variable sex differences in plasma lipid levels at different age. Our findings emphasize the importance of sex- and age-stratified analyses in lipidome studies. The study paves the way for further evaluation in sex-specific manner towards precision medicine and sex-specific genetic and lipid biomarker discovery and further highlights the need for sex-specific prevention and management of ASCVD risk.

## Materials and Methods

### Study participants

The study included participants from the following cohorts:

#### GeneRISK cohort

The study included 7,292 participants, aged 45-66 years, from the ongoing prospective GeneRISK cohort recruited during 2015–2017 from Southern Finland. The recruitment process and sample collection procedures are described in detail in [42]. Briefly, participants were instructed to fast overnight for 10 hours before the blood samples were collected for plasma, serum and DNA extraction. Fasting serum lipids including HDL-C, LDL-C, TG, TC, Apolipoprotein A1 and Apolipoprotein B were measured using standard enzymatic methods. The 10-year ASCVD risk was estimated based on clinical risk factors including age, sex, smoking status, total cholesterol (TC), HDL-C, systolic blood pressure (SBP), current usage of antihypertensive medication and family history of early-onset CHD. The ASCVD risk estimates were interpreted as outlined in the Finnish National Guidelines for prevention of coronary heart disease and/or stroke, i.e. a 10-year risk < 2% is considered low, 2-10% is intermediate, > 10% is considered high risk. GeneRISK study participants’ DNA, blood, serum, and plasma samples, in addition to their demographic information and health data have been stored in the THL Biobank (https://www.thl.fi/en/web/thlfien/topics/information-packages/thl-biobank).

#### Replication cohorts

For the replication of the findings from the lipidome analyses, we included data from 2,181 participants from the EUFAM and FINRISK cohorts with lipidomic and genetic data that we have reported previously [22]. Sample recruitment and study protocols for the EUFAM and FINRISK cohorts have been described previously [43, 44]. Briefly, EUFAM study cohort is comprised of the Finnish familial combined hyperlipidemia families. The Finnish National FINRISK study is a population-based survey conducted every 5 years since 1972, and thus far samples have been collected in 1992, 1997, 2002, 2007, and 2012, and are stored in the National Institute for Health and Welfare/THL) Biobank.

To replicate the findings from the analyses of traditional lipids, we used lipid measurements (TC, LDL-C, HDL-C, triglycerides, apolipoprotein A-1 (ApoA1) and apolipoprotein B (ApoB) for 24,614 individuals (11,238 women and 13,376 men) from the UK Biobank who had fasted between 7 to 15 hours before blood sample collection. We excluded individuals with reported fasting duration < 7 or > 15 hours to reduce potential bias that might arise due to difference in fasting duration. Information on participants’ age, BMI, diabetes, smoking status, medications and hormone replacement therapy was also obtained to be used as covariates. Details on sample handling and assays used for lipid measurements in UK Biobank cohort have been described previously [45, https://biobank.ndph.ox.ac.uk/showcase/showcase/docs/serum_biochemistry.pdf].

### Ethics statement

The study was carried out according to the principles of the Helsinki declaration and the Council of Europe’s (COE) Convention of Human Rights and Biomedicine. All study participants gave their informed consent to participate in the study. The study protocols were approved by The Hospital District of Helsinki and Uusimaa Coordinating Ethics committees (approval No. 281/13/03/00/14 (GeneRISK); approval No. 184/13/03/00/12 (EUFAM)).

### Shotgun Lipidomics

Lipidomic measurements were performed using mass spectrometry-based shotgun lipidomic analysis at Lipotype GmbH (Dresden, Germany). Samples were analyzed by direct infusion in a QExactive mass spectrometer (Thermo Scientific) equipped with a TriVersa NanoMate ion source (Advion Biosciences) [46]. Data were analyzed using in-house developed lipid identification software and developed data management system [47, 48]. Lipids with signal-to-noise ratio >5 and amounts >5-fold higher than in corresponding blank samples were considered. Reproducibility was assessed by the inclusion of 8 reference plasma samples per 96 well plate. Using 8 reference samples per 96-well plate batch, lipid amounts were corrected for batch variations and for analytical drift if the p-value of the slope was below 0.05 with an R2 greater than 0.75 and the relative drift was above 5%. Five samples with very low total lipids content and number of lipids detected were removed and lipid species detected in <70% of the remaining samples were also excluded. Furthermore, samples with >30% missingness for the QC passed lipid species were also excluded. Thus, after quality control (QC), lipidomics data in GeneRISK cohort comprised of 179 lipid species from 13 lipid classes for 7,266 individuals including 2,624 men and 4,642 women. As expected, many lipid species are highly correlated, with 70 principal components (PC) explaining >90% of the variation in the data. After the same QC procedures in the replication cohorts, data for 169 lipid species matching with GeneRISK dataset for 2,045 individuals was available.

### Genotyping and imputation

Genotyping for the GeneRISK study participants was performed using the HumanCoreExome BeadChip (Illumina Inc., San Diego, CA, USA). The genotypes were called using GenomeStudio and zCall at the Institute for Molecular Medicine Finland (FIMM). Genotyping data was lifted over to build version 38 (GRCh38/hg38) (as described in dx.doi.org/10.17504/protocols.io.nqtddwn). Pre-imputation QC included exclusion of individuals with <95% call rate, discrepancies between biological and reported sex, extreme heterozygosity (±4SD), and non-Finnish ancestry as well as of variants with <98% call rate, deviation from Hardy-Weinberg Equilibrium (HWE *P* < 1×10^−6^) and minor allele frequency (MAF) <0.05. Pre-phasing of genotyped data was performed with Eagle 2.3.5 with the number of conditioning haplotypes set to 20,000 [49]. Imputation was done with Beagle 4.1 [50] (as described in https://doi.org/10.17504/protocols.io.nmndc5e) using population-specific Sequencing Initiative Suomi (SISu) v3 reference panel developed from high-coverage (25–30x) whole-genome sequences for 3,775 Finnish individuals. Post-imputation QC included exclusion of variants with imputation info score < 0.70 and minor allele frequency < 0.01. Genotyping for both the EUFAM and FINRISK cohorts was performed using the HumanCoreExome BeadChip (Illumina Inc., San Diego, CA, USA). Details about the quality control and imputation of the EUFAM and FINRISK cohorts are described previously [22].

### Statistical analyses investigating sex differences in lipidome

All statistical analyses related to age and sex differences and data visualization were done using R 3.6.3. Associations with P < 7.1×10^−4^ (P < 0.05/70 PCs to adjust for multiple tests) were considered statistically significant. Lipid measurements were log10 transformed prior to the analyses. Age-related trends in traditional lipids, ApoA1 and ApoB were determined by calculating mean levels at each 5-year interval for age ranges 45–50-year-olds, 51–55-year-olds, 56–60-year-olds and 61–66-year-olds. To visualize changes in lipidome profiles with age, mean levels at each 1-year interval for each lipid species were calculated separately in men and women and were normalized to the respective mean levels of 45-50-year-olds (used as reference groups). For normalization, mean value of each lipid species for the reference group was subtracted from each 1-year interval mean level of that lipid and then divided by the standard deviation of the lipid in the reference group for both sexes. Linear regression analyses were performed to statistically determine relationship between lipids and age, separately in men and women, adjusted for BMI, diabetes, cardiac disease, lipid lowering medication and smoking; and additionally, for hormone replacement therapy for women. Differences between the lipidome profiles of men and women were evaluated for each 5-year interval by linear regression adjusting for age, BMI, diabetes, cardiac disease, lipid lowering medication and smoking habits. Differences between the lipidome profiles of men and women in full dataset (without age-based stratification) was additionally adjusted for age^2^. Menopause related analyses were performed in a subgroup of participants aged 45-55 years to assess age-independent effect of menopause. Menopause status was assessed through questionnaire filled at the time of recruitment and confirmed by the status reported during the follow-up visit in the GeneRISK study (∼2 years later). Only women with consistent reporting of menopause status (524 premenopausal and 663 postmenopausal women) at two visits were included. Heterogeneity in the effect sizes between two groups was estimated using the following equation:

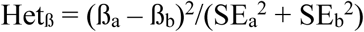

P values for heterogeneity (P_het_) were obtained from Het_ß_ under the null assumption of equal effect sizes in the two groups (referred as a and b in the equation), from the standard chi-square distribution with 1 degree of freedom.

### Sex-stratified genome-wide association analyses

For sex-stratified GWAS in the GeneRISK cohort, residuals obtained after regressing for age, age^2^, collection site, lipid medication, first 10 genetic PCs; and additionally for menopause status and hormone replacement therapy in women, were inverse-normal transformed separately in men and women and were used as outcome variables. GWASs were performed using linear regression model implemented in Plink2.0 [http://pngu.mgh.harvard.edu/purcell/plink/, 51]. Heterogeneity in the effect sizes between men and women was assessed using the equation provided above for Het_ß_. In the genome-wide scans, associations with P < 7.1×10^−10^ (5.0×10^−8^/70 PCs explaining >90% variance) were considered statistically significant. For the replication of the identified associations, association analyses in the replication cohort were performed using sex-specific inverse-normal transformed lipid levels adjusted for age, age^2^, cohort, lipid medication, familial hyperlipidemia, first 10 genetic PCs, and additionally for hormone replacement therapy in women using linear mixed model implemented in MMM [52].

## Data Availability

The summary level data and raw data underlying the main text and figures are contained in Supplementary Data. The UK Biobank resource is available to bona fide researchers for health-related research in the public interest at https://www.ukbiobank.ac.uk/researchers/. Other data are available through the Institute for Molecular Medicine Finland Data Access Committee on reasonable request after appropriate ethical approval.

## Code availability

The full genotyping and imputation protocol is described in https://doi.org/10.17504/protocols.io.nqtddwn All software packages and programs used to perform these analyses are freely available. The code used for these analyses are available from the corresponding authors upon reasonable request.

## Acknowledgements

This research has been conducted using the UK Biobank Resource under Application Number 22627. We would like to thank Johanna Aro, Sari Kivikko, Ulla Tuomainen for management assistance in the project.

---

**S1 Fig:**
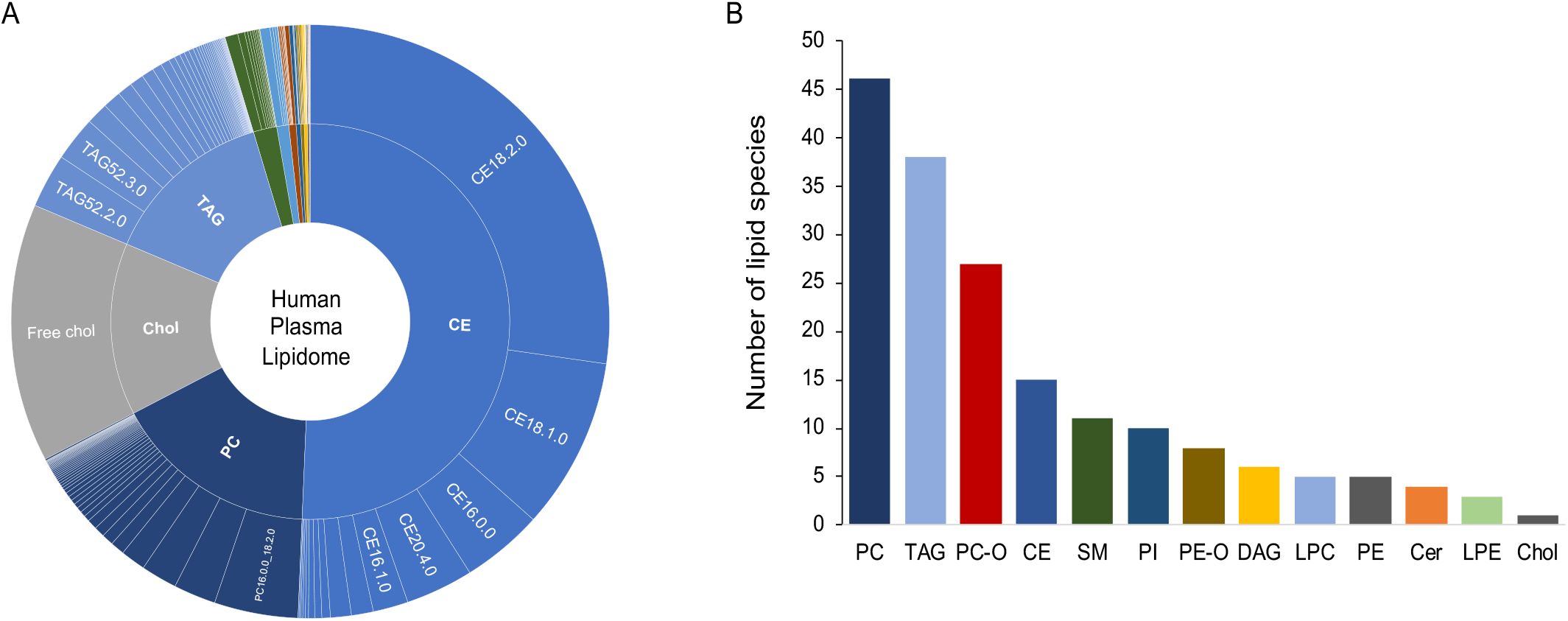
Details of lipid species detected in the GeneRISK cohort. **(A) Relative abundance of lipid species in human plasma lipidome**. The outer circle shows the proportion of each lipid species in human plasma as determined by the mean plasma level of lipid species divided by the mean total lipids (sum of all 179 lipids detected in our dataset). The inner circle shows the proportion of a lipid class in human plasma determined as the mean of sum of all lipid species in a lipid class divided by the mean total lipids. **(B) Number of lipid species detected in each lipid class** - CE: Cholesteryl esters; Cer: Ceramides; DAG: Diacylglycerides; LPC: Lysophosphatidylcholines; LPE: Lysophosphatidylamines; PC: Phosphatidylcholines; PC-O: Phosphatidylcholine-ethers; PE: Phosphatidylamines, PE-O: Phosphatidylamine-ethers; PI: Phosphatidylinositols; SM: Sphingomyelins; TG: Triacylglycerides; Chol: Free cholesterol. The lipid classes are represented by the same colors in A and B.

**S2 Fig:**
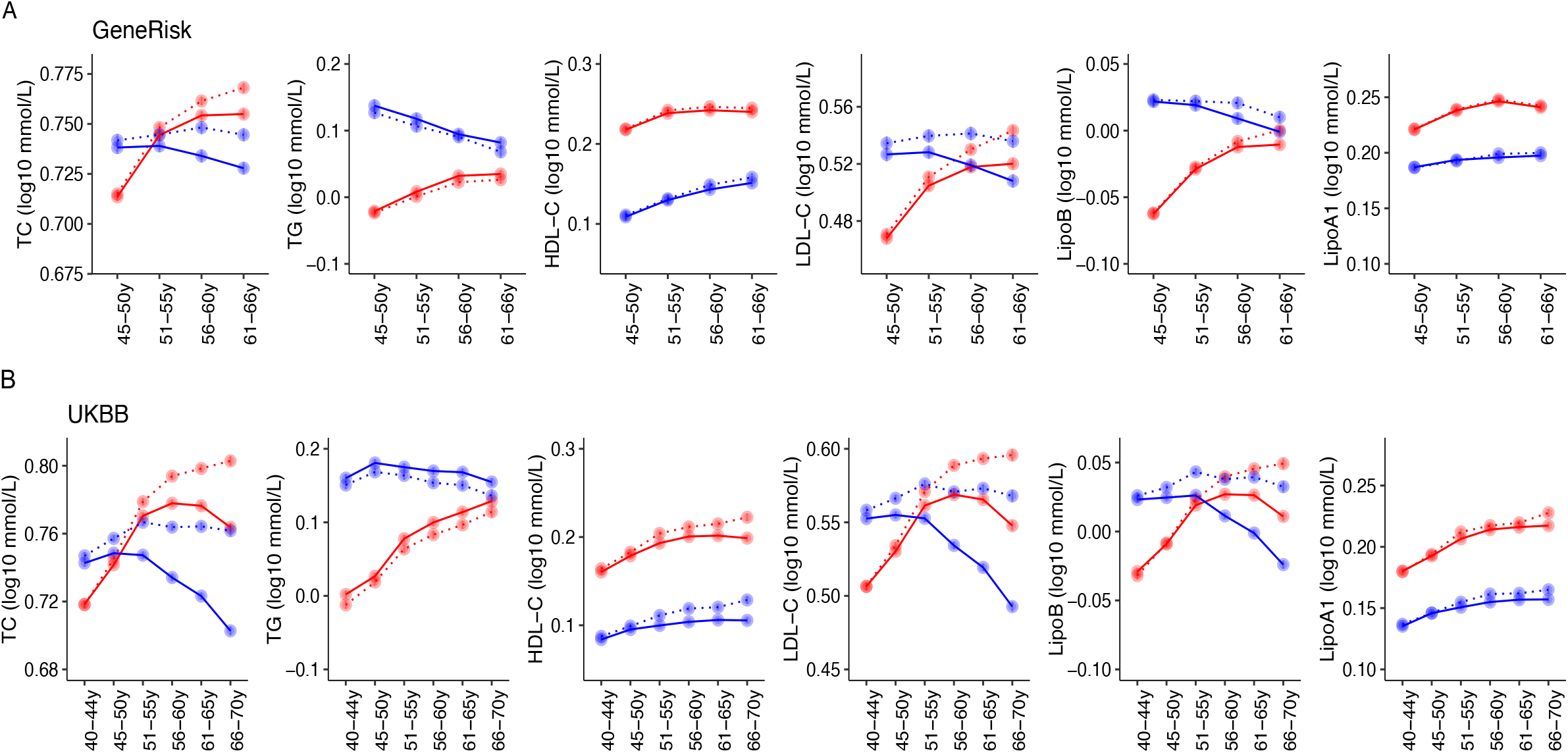
Age-related trends in traditional lipid in GeneRISK (A) and UKBB (B). Mean levels after log10 transformation and standard errors of total cholesterol, triglycerides, HDL-C, LDL-C, Apolipoprotein B and Apolipoprotein A-1 at 5-year intervals are plotted for men (blue solid lines) and women (red solid lines). Dotted lines present the trends for lipid levels after excluding the individuals with lipid lowering medication from the analyses.

**S3 Fig:**
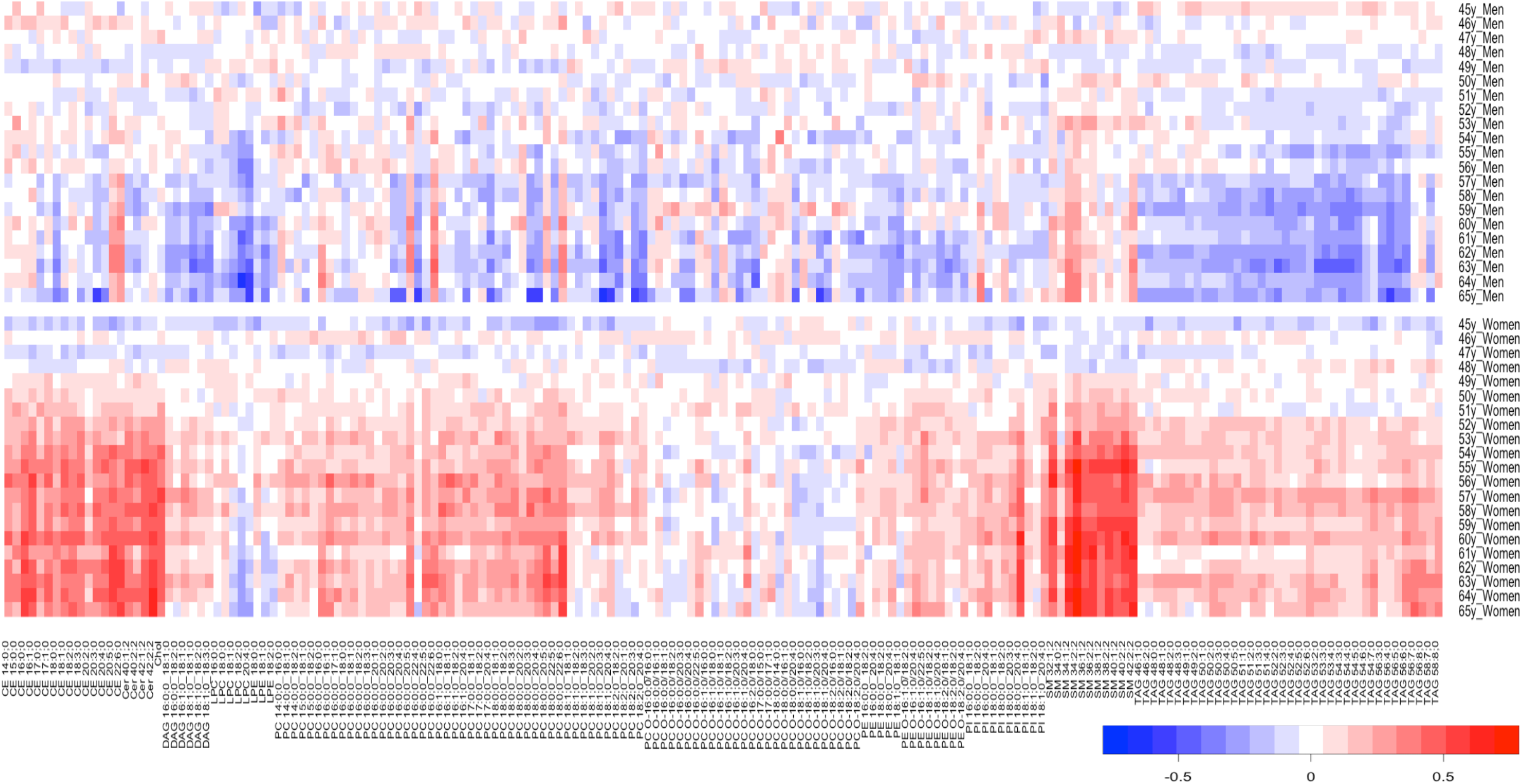
Age-related trends in lipidome profiles. Change in molecular lipid species across 1-year intervals in men (upper panel) and women (lower panel). The average lipid species levels (adjusted for covariates) were calculated for each 1-year interval and centered and scaled to the mean and standard deviation of the reference age-group 44-50-years.

**S4 Fig:**
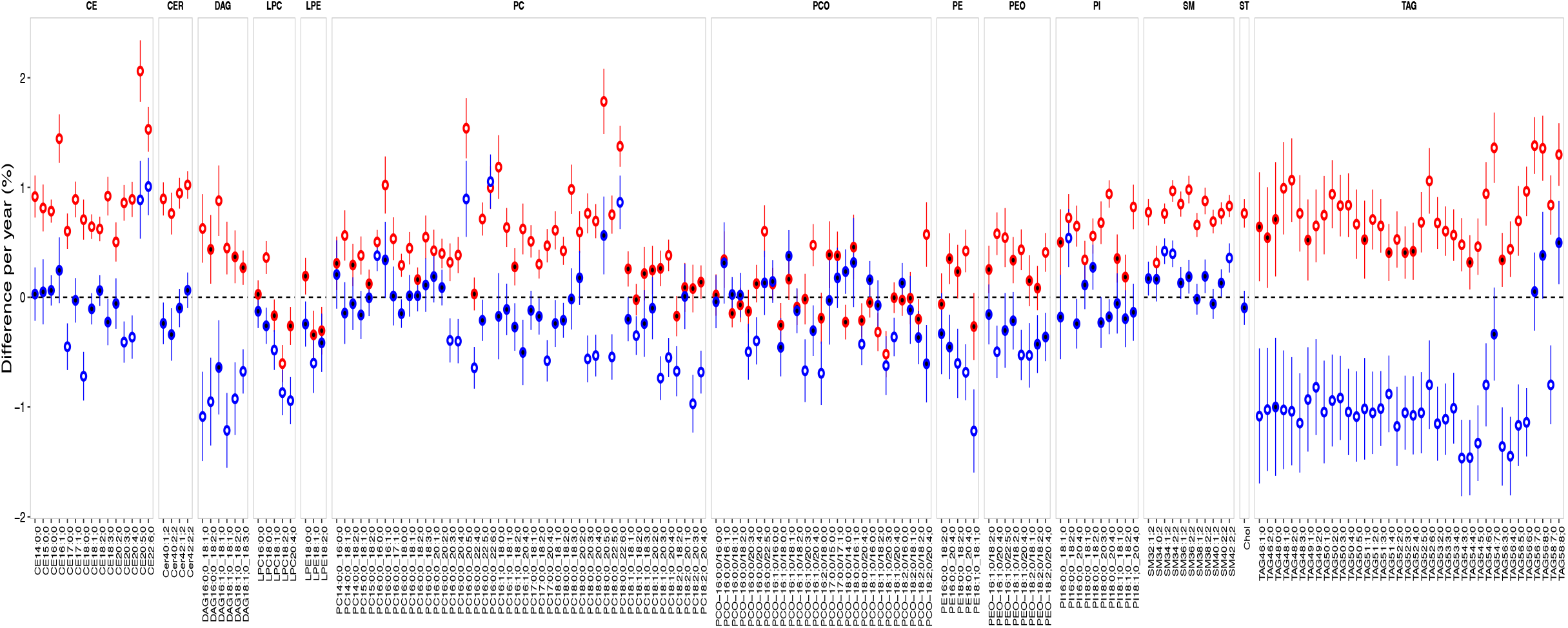
Association of age with lipid species in the GeneRISK cohort. Percent change in lipid levels (log10 transformed) per year for men (blue) and women (red) are plotted on y-axis. The error bars represent the 95% confidence interval. The significant associations are filled with white color. The lipid species are grouped by their lipid classes and are arranged in increasing number of carbon atoms and double bonds. The percent differences were calculated from beta coefficients obtained from linear regression analyses with lipid levels as dependent variable and age as independent variables along with covariates mentioned in the methods (difference (%) =100 x (10^β^-1)).

**S5 Fig:**
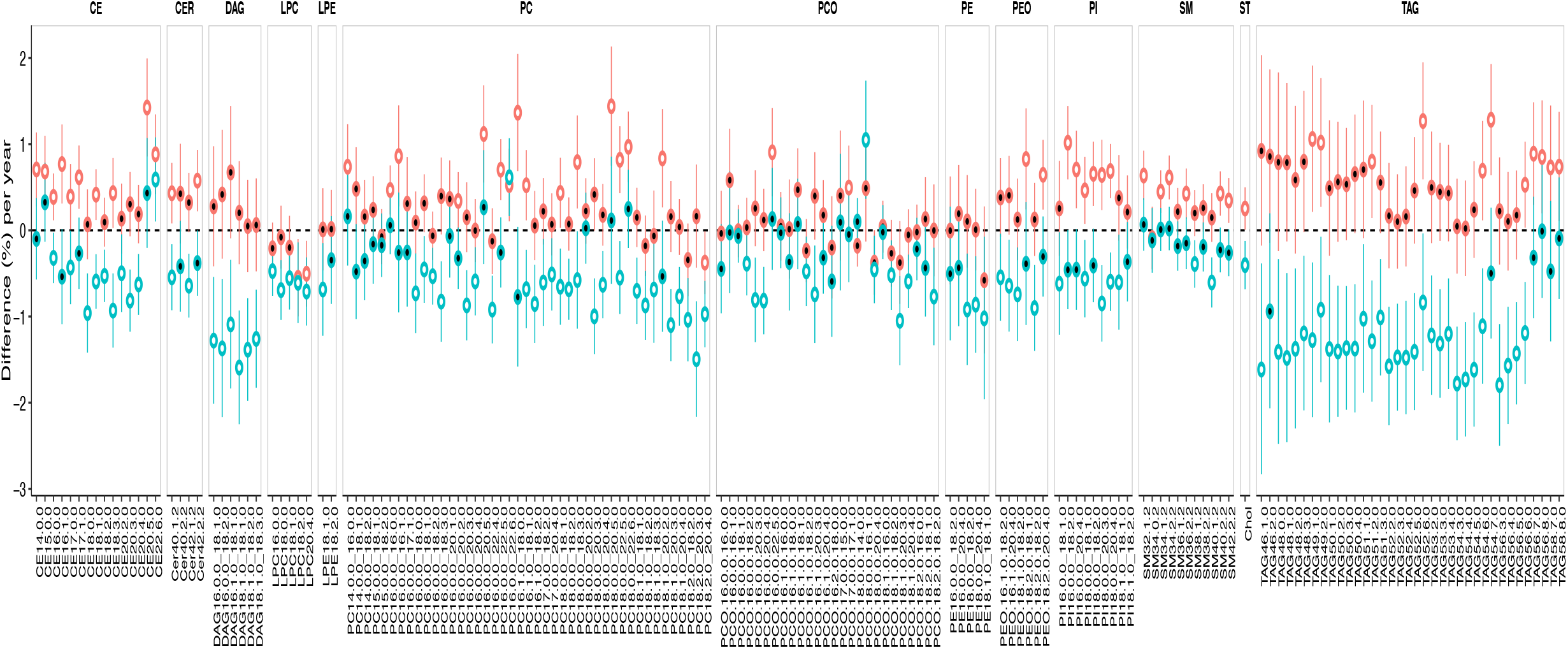
Association of age with lipidome in replication cohort. Percent change in lipid levels (log10 transformed) per year for men (blue) and women (red) are plotted on y-axis. The error bars represent the 95% confidence interval. The significant associations are filled with white color.

**S6 Fig:**
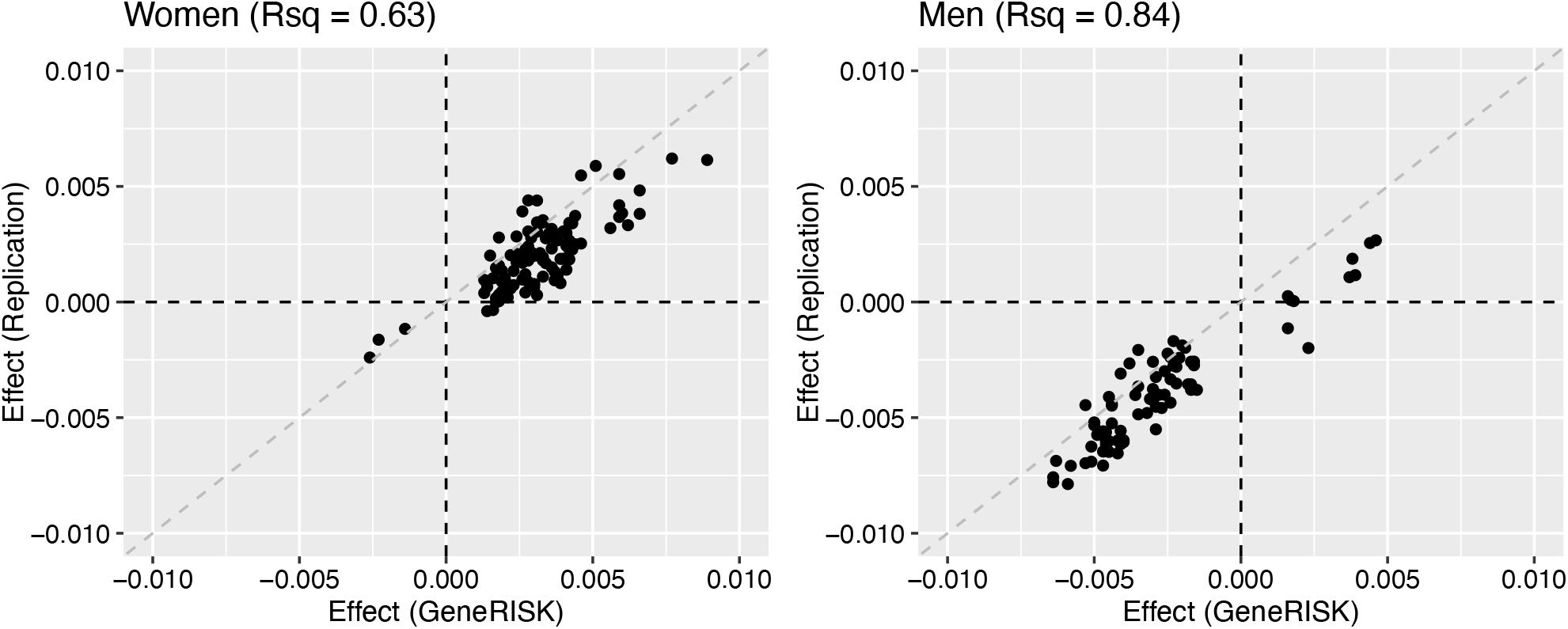
Comparison of effect size for association with age in the GeneRISK and the replication cohort. Lipid species with significant association with age in the GeneRISK cohort are plotted.

**S7 Fig:**
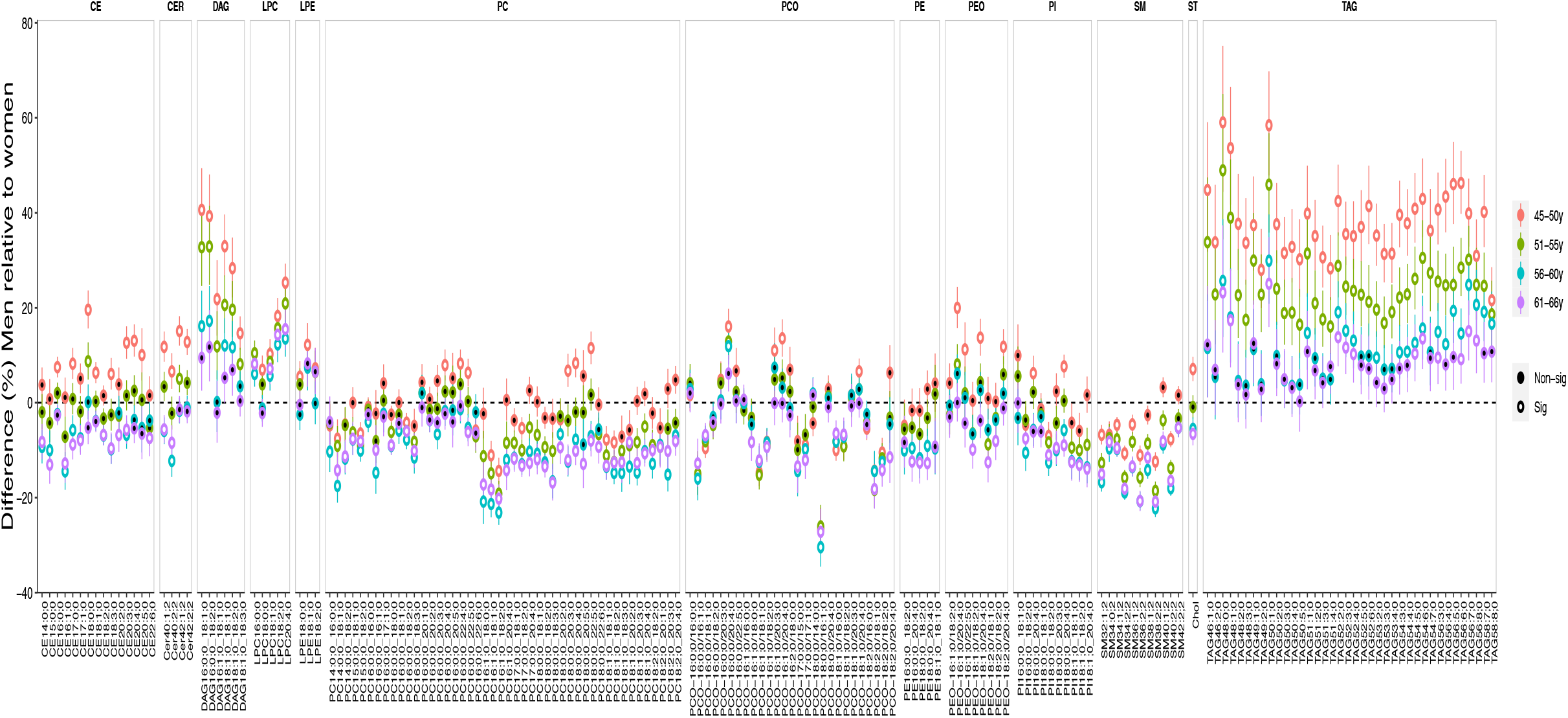
Age-dependent sex differences in plasma lipidome in the GeneRISK cohort. Difference in effect of age on lipid levels (percent) in men relative to women determined separately for age-groups 45-50 years (red), 51-55 years (green), 56-60 years (blue) and 61-66 years (purple) along with their 95% confidence intervals are plotted on y-axis. The effects were calculated from beta coefficients of male sex obtained from linear regression analyses with lipid levels as dependent variable and male sex as independent variables along with covariates mentioned in the methods and turned into percentages (difference (%) =100 x (10^β^-1)). The error bars represent the 95% confidence interval. The significant associations in linear regression models adjusted for covariates (P < 7.0×10^−4^) are filled with white color.

**S8 Fig:**
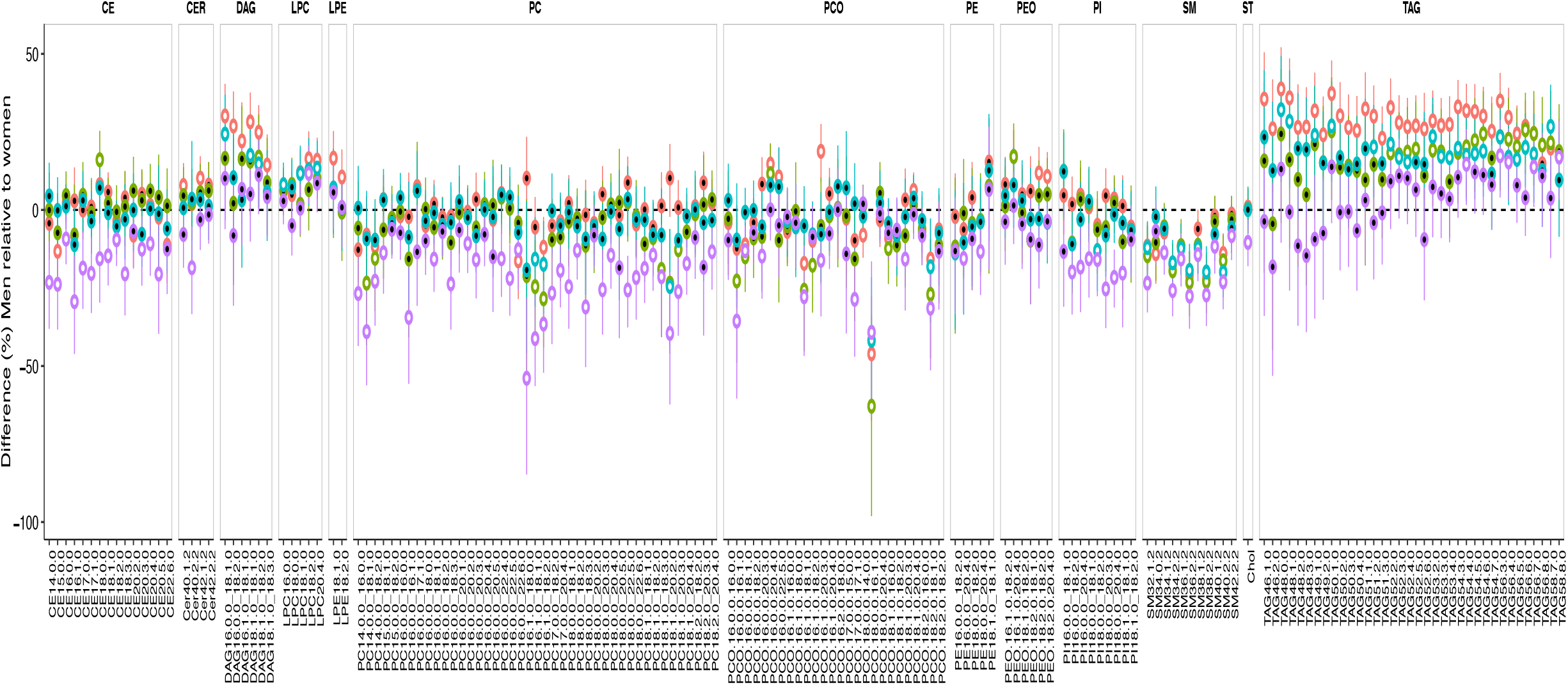
Age-dependent sex differences in plasma lipidome in the replication cohort. Percent difference in lipid levels (log10 transformed) in men relative to women are plotted on y-axis for age-groups - 45-50 years (red), 51-55 years (blue), 56-60 years (green) and 61-66 years (purple). The percent differences were calculated from beta coefficients obtained from linear regression analyses with lipid levels as dependent variable and age as independent variables along with covariates mentioned in the methods (difference (%) =100 x (10^β^-1)). The error bars represent the 95% confidence interval. The significant associations in linear regression models adjusted for covariates are filled with white color.

**S9 Fig:**
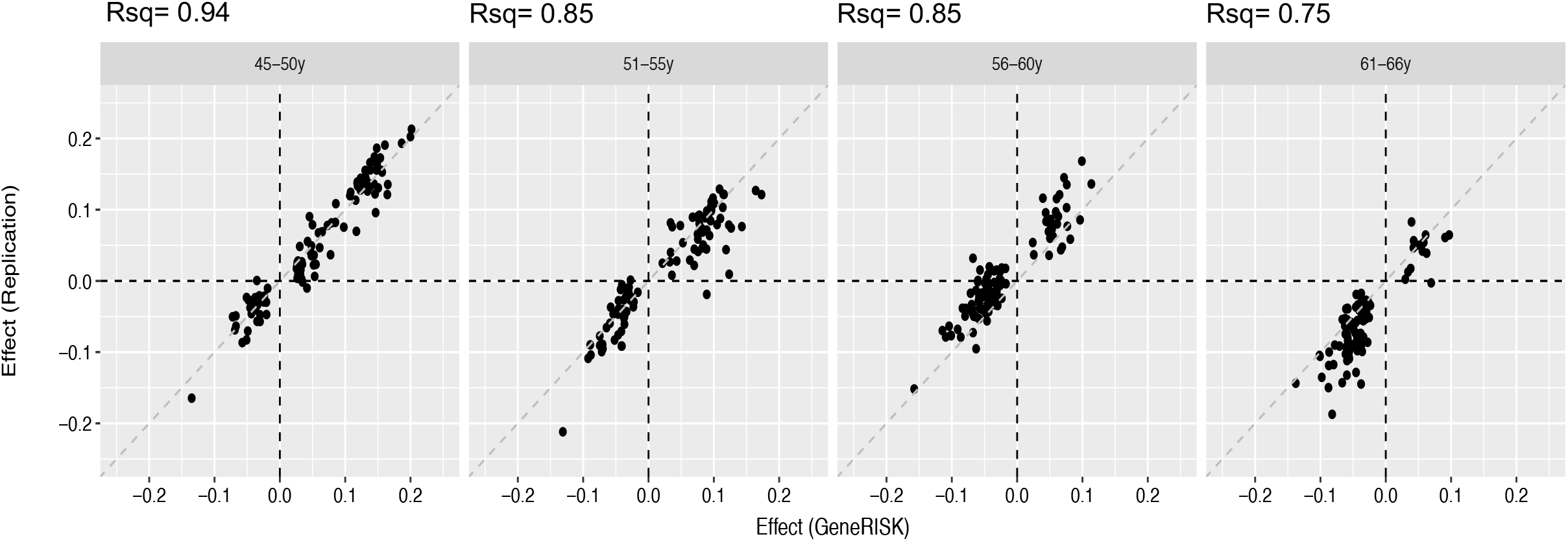
Comparison of effect size for association with age in the GeneRISK and replication cohort. The effect sizes for association of age on different lipid species are plotted on x-axes for the GeneRISK cohort and on y-axes for the replication cohort in different age groups.

**S10 Fig:**
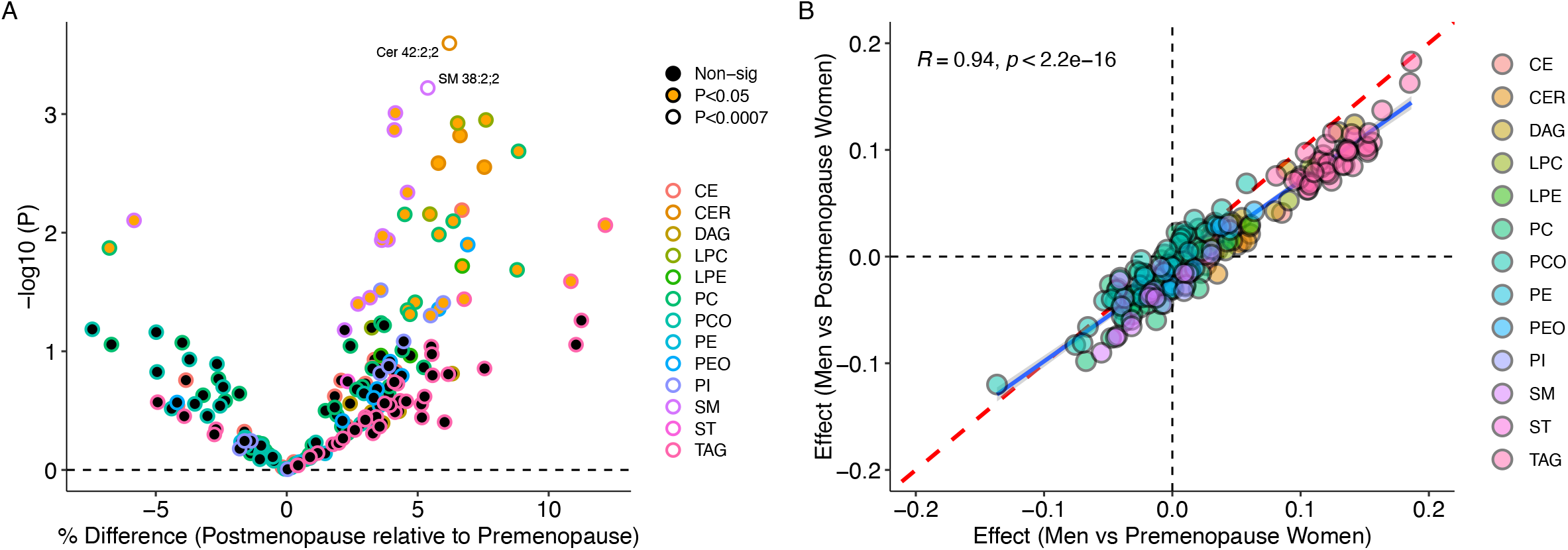
Effect of menopause on sex differences in lipidome. (A) Association between menopause and levels of lipid species. The percent difference in postmenopausal women relative to premenopausal women are plotted on x-axis with corresponding P values from linear regression analysis on y-axis. (B) Scatter plot showing comparison between the effect sizes for association with male gender in dataset including only premenopausal women (x-axis) and effect sizes for association with male gender in dataset including only postmenopausal women (y-axis).

**S11 Fig:**
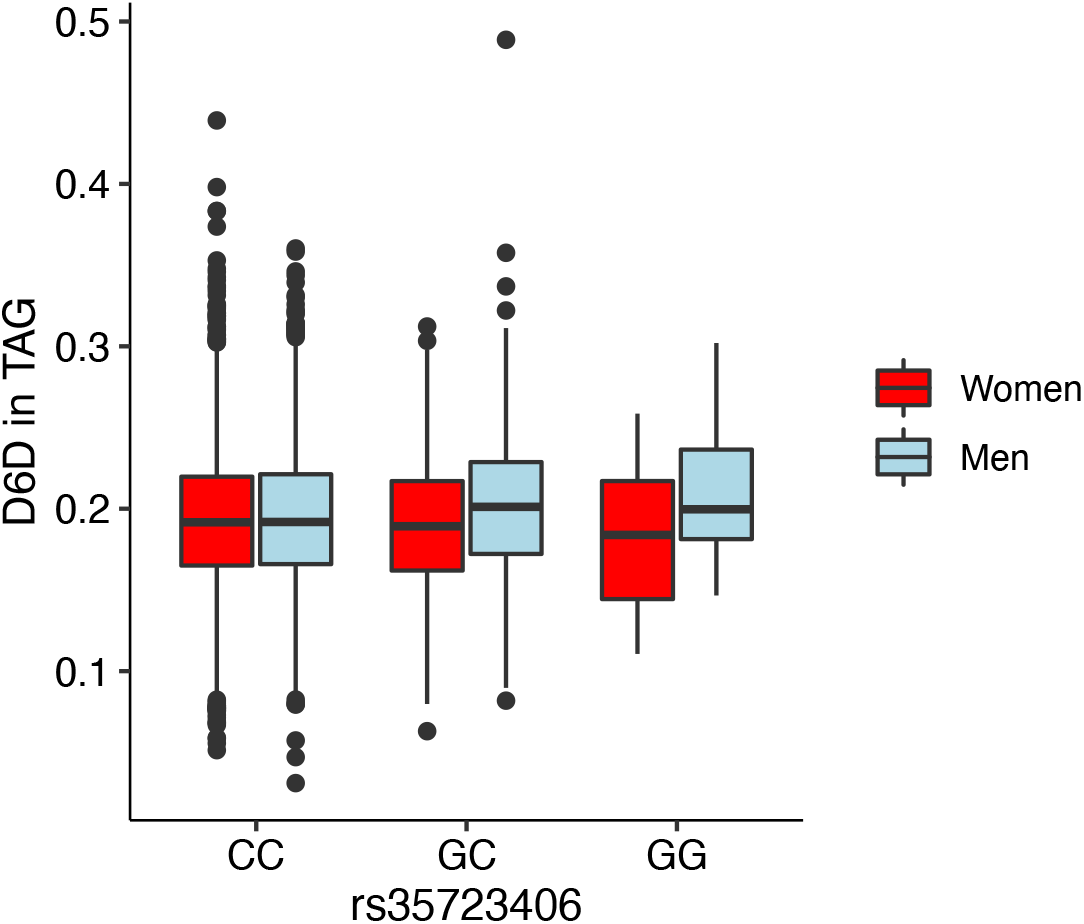
Association of rs35723406 in *TKFC* gene near FADS1-2-3 region with lipid index estimate for Δ6 desaturase in TAG (D6D in TAG). The boxplot shows the levels of D5D in TAG in men and women with different genotypes.

